# Different selection dynamics of S and RdRp between SARS-CoV-2 genomes with and without the dominant mutations

**DOI:** 10.1101/2021.01.03.20237602

**Authors:** Necla Koçhan, Doğa Eskier, Aslı Suner, Gökhan Karakülah, Yavuz Oktay

## Abstract

SARS-CoV-2 is a betacoronavirus responsible for the COVID-19 pandemic that has affected millions of people worldwide, with no dedicated treatment or vaccine currently available. As pharmaceutical research against and the most frequently used tests for SARS-CoV-2 infection both depend on the genomic and peptide sequences of the virus for their efficacy, understanding the mutation rates and content of the virus is critical. Two key proteins for SARS-CoV-2 infection and replication are the S protein, responsible for viral entry into the cells, and RdRp, the RNA polymerase responsible for replicating the viral genome. Due to their roles in the viral cycle, these proteins are crucial for the fitness and infectiousness of the virus. Our previous findings had shown that the two most frequently observed mutations in the SARS-CoV-2 genome, 14408C>T in the RdRp coding region, and 23403A>G in the S gene, are correlated with higher mutation density over time. In this study, we further detail the selection dynamics and the mutation rates of SARS-CoV-2 genes, comparing them between isolates carrying both mutations, and isolates carrying neither. We find that the S gene and the RdRp coding region show the highest variance between the genotypes, and their selection dynamics contrast each other over time. The S gene displays higher positive selection in mutant isolates early on, and undergoes increasing negative selection over time, whereas the RdRp region in the mutant isolates shows strong negative selection throughout the pandemic.

## 1. Introduction

COVID-19 is a pandemic characterized caused by the SARS-CoV-2 betacoronavirus, of zoonotic origin, with human-to-human transmission capacity, and since the first observed cases in the Wuhan province of China (Chan et al., 2020; Riou & Althaus, 2020), it has infected 45,968,799, with over 1 million recorded deaths (as of 6 November 2020), with 69% of cases and 78% of deaths in Europe and the Americas. While its primary characteristics are respiratory system distress and long-onset fever, long term symptoms include neuroinvasion (Li, Bai & Hashikawa, 2020; Wu et al., 2020), cardiovascular complications (Kochi et al., 2020; Zhu et al., 2020), and gastrointestinal and liver damage (Lee, Huo & Huang, 2020; Xu et al., 2020). Alongside these secondary symptoms, the disease is also capable of asymptomatic community-wide transmission due to its high transmissibility (Wong et al., 2020), creating a unique global health risk of urgent interest. As a result, the high amount of frequently updated data on viral genomes on databases such as GISAID (Elbe & Buckland-Merrett, 2017) and NextStrain (Hadfield et al., 2018) provides researchers with invaluable resources to track the evolution of the virus.

SARS-CoV-2 has a linear, single-stranded genome, and encodes its own genomic replication proteins, instead of relying on host proteins. The replication proteins are cleaved from the Orf1ab polyprotein, and the most critical among them, RdRp (RNA-dependent RNA polymerase), is responsible for synthesizing new strands of RNA using the viral genome as a template, which together with nsp7 and nsp8 creates the core polymerase complex (Kirchdoerfer & Ward, 2019; Peng et al., 2020), while nsp14 is an exonuclease which provides error-correcting capability to the RNA synthesis complex, therefore allowing the SARS-CoV-2 to maintain its large size genome (Subissi et al., 2014; Ma et al., 2015; Ogando et al., 2019; Romano et al., 2020). Owing to their role in maintaining replication fidelity and directly affecting the mutation-selection equilibrium of RNA viruses, these proteins are key targets of study in understanding the mutation accumulation and adaptive evolution of the virus (Eckerle et al., 2010; Peng et al., 2020).

One of the key mutations in the SARS-CoV-2 RdRp coding region is the 14408C>T mutation, frequently co-occurring with the 23403A>G mutation of the S gene. In our previous study, we examined the top 10 most frequent mutations in the SARS-CoV-2 RdRp, and identified that four of them are associated with an increase in mutation density in two genes, the membrane glycoprotein (M) and the envelope glycoprotein (E) which are under relatively low selective pressure, and mutations in these genes can be used to infer reduced replication fidelity (Eskier et al., 2020a). The mutation that most strongly affects the mutation density was the 14408C>T mutation, predicating it as a genotype of interest. In our follow-up study, we also identified the trend of a positive correlation between the presence of the 14408C>T mutation and increased mutation density in regions under low selective pressure persisted beyond our initial study (Eskier et al., 2020c).

## 2. Material & Methods

### Genome sequencing and preprocessing

As previously described (Eskier et al., 2020a), SARS-CoV-2 isolate genome sequences from United Kingdom and the United States, and the corresponding metadata, were obtained from the GISAID EpiCoV database (date of accession: 7 September 2020). We applied further quality filters, including selecting only isolates obtained from human hosts (excluding environmental samples and animal hosts), those sequenced for the full length of the genome (sequence size of 29 kb or greater), and those with high coverage for the reference genome (< 1% N content, < 0.05% unique mutations, no unverified indel mutations). To ensure alignment accuracy, all nonstandard unverified nucleotide masking was changed to N due to the specifications of the alignment software, using the Linux *sed* command, and the isolates were aligned against the SARS-CoV-2 reference genome (NCBI Reference Sequence NC_045512.2, available at https://www.ncbi.nlm.nih.gov/nuccore/NC_045512.2) using the MAFFT (v7.450) alignment software (Katoh et al., 2002), using the parameters outlined in the software manual for aligning closely related viral genomes (available at https://mafft.cbrc.jp/alignment/software/closelyrelatedviralgenomes.html). Aggregate country-specific FASTA files were each divided into the reference sequence plus 1000 isolate genomes using the fasta-splitter software (version 0.2.6, available at http://kirill-kryukov.com/study/tools/fasta-splitter/). The FASTA files were converted into PHYLIP files for downstream analyses using the msa_view utility of the PHAST software package (last updated 2 July 2019), with the parameters “-i FASTA -o PHYLIP input.file > output.file” (Hubisz, Pollard & Siepel, 2011). Following all filters, a total of 34,393 genomes were used in the study (18,125 for UK, 16,268 for the US). We further divided the remaining isolate sequences into mutant (MT) and wildtype (WT) genotypes, with isolates carrying the 14408C>T and 23403A>G mutations classified as MT, while isolates carrying neither mutation classified as MT.

### Modeling the daily mutation rate

In this study, we considered the conservative constant mutation rate model (Vega et al., 2004), given as follows:

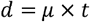

where, the number of mutations (*d*) occurred in an isolate from its ancestor is calculated as the multiplication of the daily mutation rate (*µ*) and the time difference (*t*) between the isolate and the NC_045512.2 reference genome. Based on the reference genome, we calculated the number of mutations in each isolate and the time difference between the isolate and the NC_045512.2 reference genome. We then estimated the daily mutation rates using least square fitting and measured the goodness of fit using the adjusted R-square statistic.

### Evolutionary rate assessment

The rate of accumulation of synonymous mutations per synonymous site (dS), the rate of accumulation of non-synonymous mutations per non-synonymous site (dN) and their ratio (dN/dS), which is also known as *w* or *K*_*a*_ /*K*_*S*_ were calculated using PAML package (version 4.9j, available at http://abacus.gene.ucl.ac.uk/software/paml.html) (Yang, 2007), with the parameters “runmode=-2, Codonfreq=2” in codeml.ctl (pairwise sequence comparison).

### Statistical Analysis

All statistical analyses were performed using the R statistical computing environment (v.4.0.2). Kolmogorov–Smirnov test was used to check the normality assumption of the continuous variables. The Wilcoxon rank-sum (Mann-Whitney U) test was performed to determine whether the difference between the WT and MT groups was statistically significant for each gene in terms of *dN, dS* and the *dN/dS* ratio. Kruskal Wallis test was performed to test if there is any significant difference between dN, dS and dN/dS of each gene in different time intervals. When there is a significant difference between time intervals, we performed multiple pairwise comparisons between time intervals using Wilcoxon rank-sum test to investigate which time intervals are statistically significant. A *p*-value less than 0.05 was considered statistically significant in all hypothesis tests.

## 3. Theory

In order to better understand the impact of the mutant genotype (hereafter referred to as MT) with 14408C>T and 23403A>G mutations on SARS-CoV-2 genome evolution and mutation rates, we used standard mathematical evolutionary biology models (Nei, Kumar & Nei, 2000; Vega et al., 2004). Our results show that evolutionary forces affect mutant genomes differently compared to genomes without these two mutations. Specifically, the S gene and the RdRp coding region seem to be the most differentially affected, albeit in opposite directions.

## 4. Results

### The effects of selective pressures are most evident in S and RdRp

In this study, we focused on the mutational dynamics of SARS-CoV-2 in UK and US populations, two regions with the highest number of isolate sequences submitted to GISAID. In particular, as our previous findings pointed to differences in mutation profiles between the two groups, we compared isolates that carried the 14408C>T and the 23403A>G mutations to isolates that did not. Isolates that carried both mutations were categorized as “mutant” (MT) while the isolates that carried neither mutation were categorized as “wild-type” (WT). Isolates that carried one mutation but not the other were removed from the analysis. We therefore divided the isolates into the following categories: UK-WT (3,412 genomes), UK-MT (14,713 genomes), US-WT (2,839 genomes) and US-MT (13,428 genomes).

In order to identify how the mutation rates of the SARS-CoV-2 genes vary, we calculated the synonymous and non-synonymous mutation rates (dS and dN, respectively) for each gene and each country (UK and US) separately (Figure 1). Our results show that the non-synonymous mutation rates are lower than the synonymous mutation rates for all genes except E. This indicates that most genes have been more strongly affected by negative selection than positive selection.

**Figure 1.**
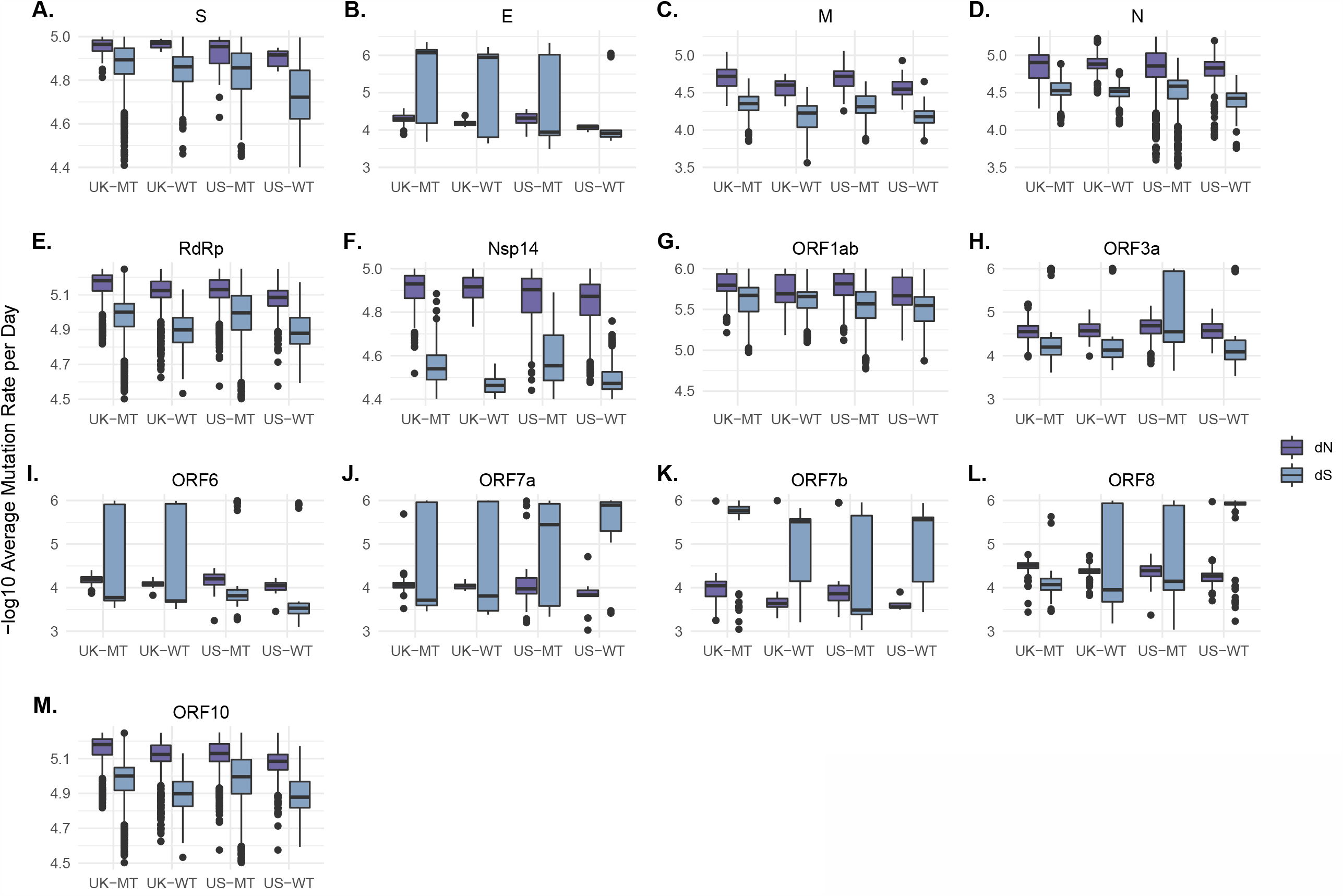
Gene wide average dN and dS values per day for country - phenotype pairings of the SARS-CoV-2 genome. (A-M) Box-and-whiskers plots for mutation rates for the genes or coding regions for the S protein (A), the E protein (B), the M protein (C), the N protein (D), the RdRp (E) and nsp14 (F) coding regions of the ORF1ab gene, the ORF1ab polyprotein (G), the ORF3a protein (H), the ORF6 protein (I), the ORF7a protein (J), the ORF7b protein (K), the ORF8 protein (L), and the ORF10 protein (M). Due to the -log10 scale, higher values on the y-axis indicate lower mutation rates.

To infer the magnitude of selection imbalance, we calculated the dN/dS ratio per day for each gene (Figure 2). The dN/dS ratios for E, M, Nsp14, ORF6, ORF7a in MT and WT isolates do not display high disparity in either country. The two genomic regions with the highest disparity were the RdRp coding region and the S genes, two regions carrying the MT-genotype defining mutations. Moreover, while the dN/dS ratio decreased for the S gene in MT isolates in both countries, it increased for the RdRp gene in the same isolates, whereas the dN/dS ratio for the ORF7b and ORF8 genes show a significant increase in US-MT isolates compared to US-WT isolates, while a similar increase is not observed in the UK isolates. We also performed the Wilcoxon rank-sum test to investigate the difference between the genotypes for individual genes in each country (Table 1). The results show a significant difference in dN, dS and the dN/dS ratio between WT and MT groups for both countries in the RdRp coding region and the S gene (p<0.001). These results also corroborate previous findings on S and RdRp, and underline the importance of these two proteins for SARS-CoV-2 evolution and fitness.

**Table 1.**
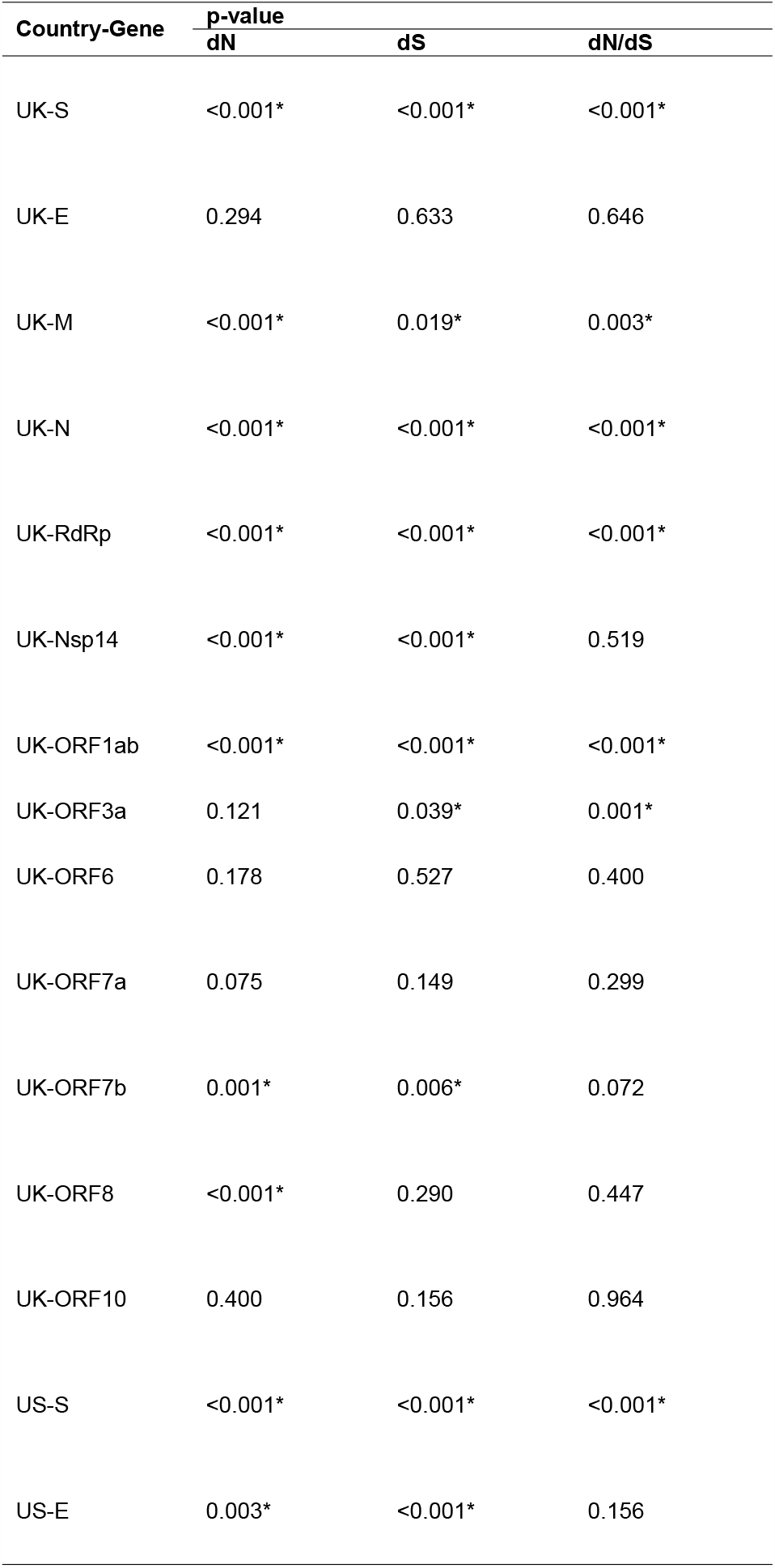
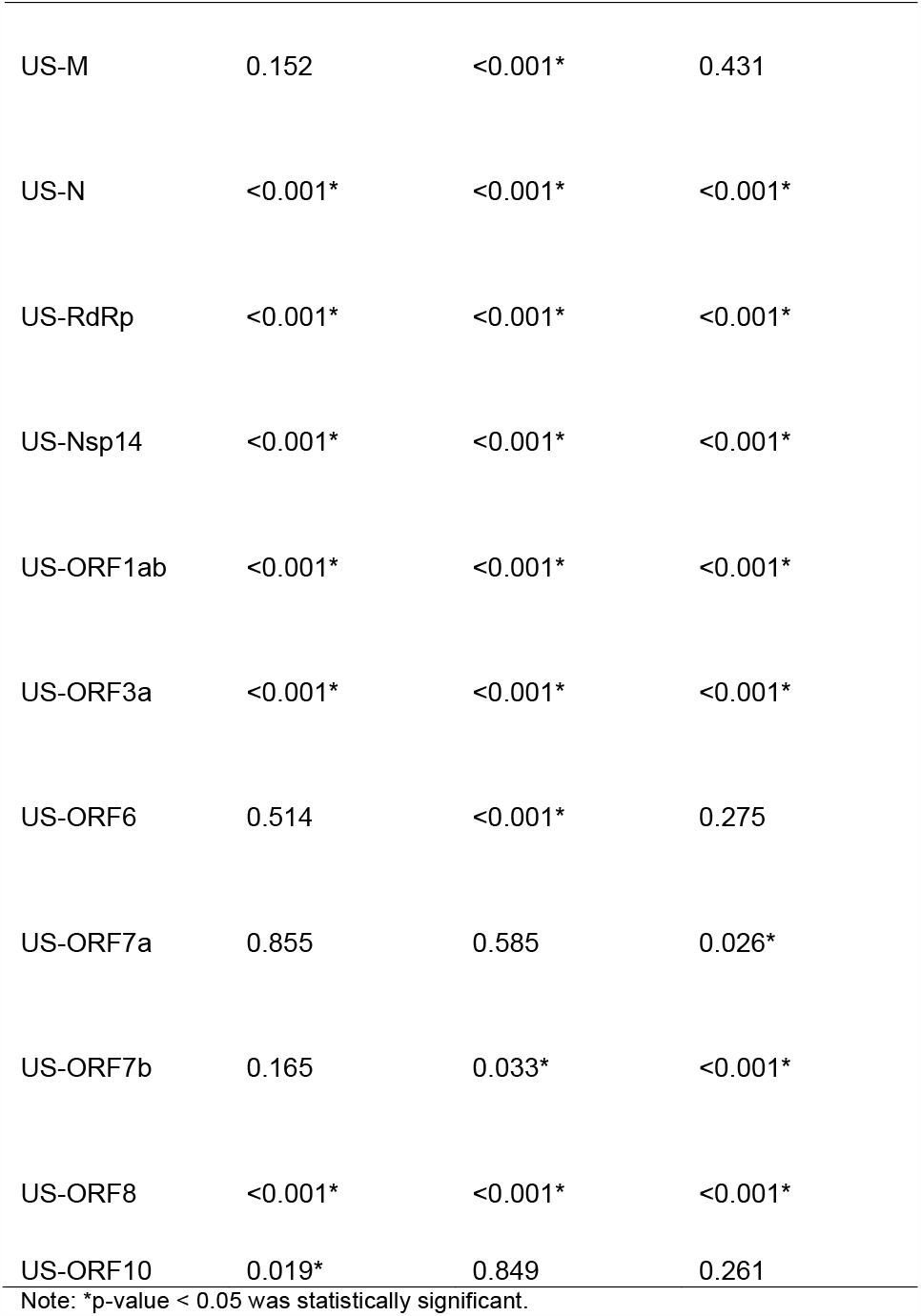
Comparison of WT and MT groups in UK and US for different coding regions

| Country-Gene | p-value |  |  |
| --- | --- | --- | --- |
|  | dN | dS | dN/dS |
| UK-S | <0.001* | <0.001* | <0.001* |
| UK-E | 0.294 | 0.633 | 0.646 |
| UK-M | <0.001* | 0.019* | 0.003* |
| UK-N | <0.001* | <0.001* | <0.001* |
| UK-RdRp | <0.001* | <0.001* | <0.001* |
| UK-Nsp14 | <0.001* | <0.001* | 0.519 |
| UK-ORF1ab | <0.001* | <0.001* | <0.001* |
| UK-ORF3a | 0.121 | 0.039* | 0.001* |
| UK-ORF6 | 0.178 | 0.527 | 0.400 |
| UK-ORF7a | 0.075 | 0.149 | 0.299 |
| UK-ORF7b | 0.001* | 0.006* | 0.072 |
| UK-ORF8 | <0.001* | 0.290 | 0.447 |
| UK-ORF10 | 0.400 | 0.156 | 0.964 |
| US-S | <0.001* | <0.001* | <0.001* |
| US-E | 0.003* | <0.001* | 0.156 |
| US-M | 0.152 | <0.001* | 0.431 |
| US-N | <0.001* | <0.001* | <0.001* |
| US-RdRp | <0.001* | <0.001* | <0.001* |
| US-Nsp14 | <0.001* | <0.001* | <0.001* |
| US-ORF1ab | <0.001* | <0.001* | <0.001* |
| US-ORF3a | <0.001* | <0.001* | <0.001* |
| US-ORF6 | 0.514 | <0.001* | 0.275 |
| US-ORF7a | 0.855 | 0.585 | 0.026* |
| US-ORF7b | 0.165 | 0.033* | <0.001* |
| US-ORF8 | <0.001* | <0.001* | <0.001* |
| US-ORF10 | 0.019* | 0.849 | 0.261 |

Note: \*p-value < 0.05 was statistically significant.

**Figure 2.**
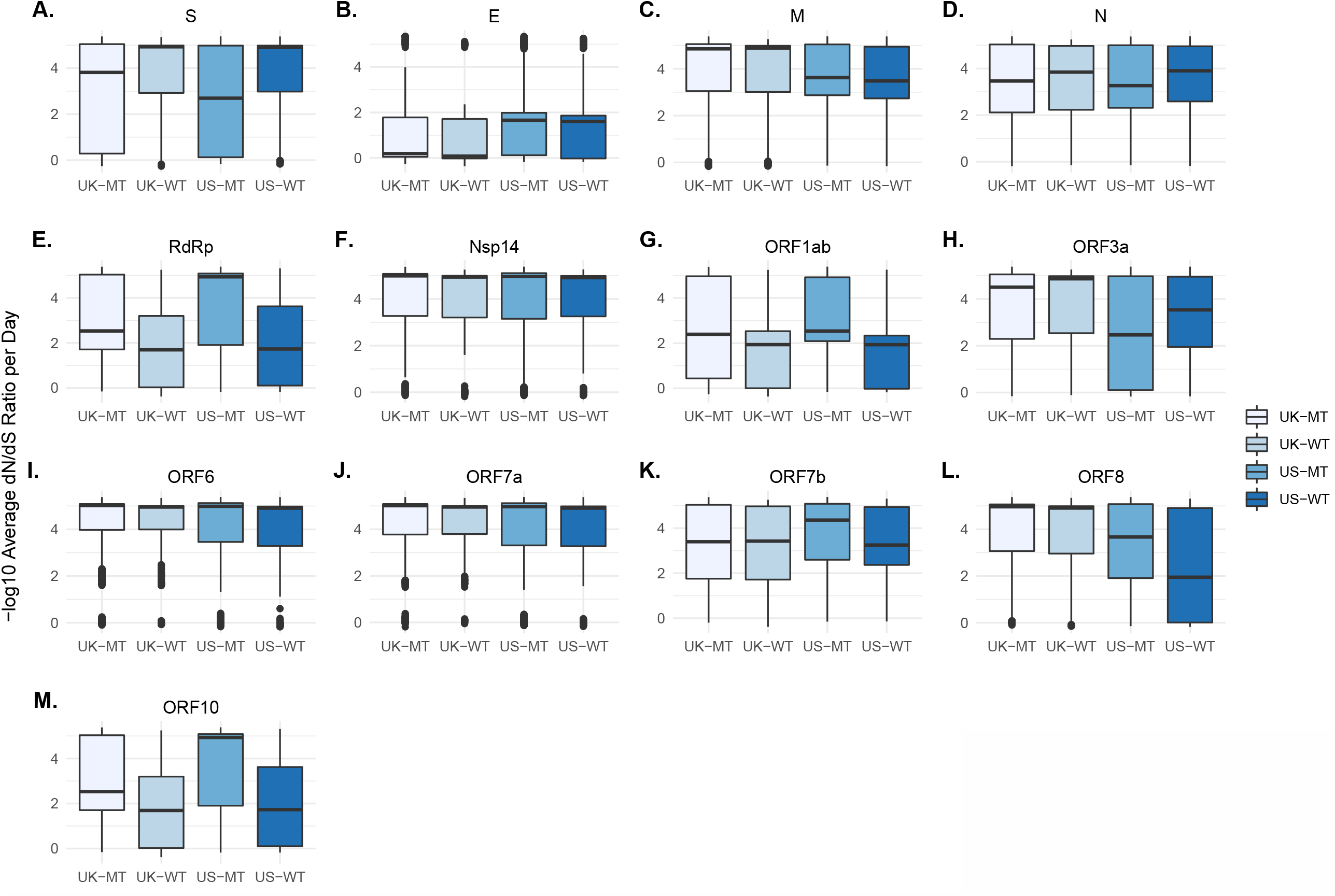
Gene wide dN/dS ratios per day for country - phenotype pairings of the SARS-CoV-2 genome. (A-M) Box-and-whiskers plots for dN/dS ratios for the genes or coding regions for the S protein (A), the E protein (B), the M protein (C), the N protein (D), the RdRp (E) and nsp14 (F) coding regions of the ORF1ab gene, the ORF1ab polyprotein (G), the ORF3a protein (H), the ORF6 protein (I), the ORF7a protein (J), the ORF7b protein (K), the ORF8 protein (L), and the ORF10 protein (M). Due to the -log10 scale, higher values on the y-axis indicate lower mutation rates.

### Stages of pandemic and viral distribution affect selective pressures and mutation rates

Our previous findings had shown a shift in terms of mutation density increase between two time categories (days 60-100 and days 101-140) in the UK and the US, as day 100 (2 April 2020) marked a turning point and a plateau in the number of daily cases in both regions. Interestingly, the mutation density averages per day were parallel to the daily case numbers and plateaued after day 100 in WT isolates, whereas it was evident in the MT isolates much later, i.e. after day 120 (Eskier et al., 2020b). Since we have witnessed similar daily case number trends between days 174-203 and days 204-242 in the US, we tested whether the mutation rates show similar patterns, as well. Therefore, we divided the US isolates into four time categories that reflect the differing dynamics of SARS-CoV-2 infection across the US population: days 80-99, days 100-143, days 174-203, and days 204-242 (Figure 3). Our results revealed that both the RdRp coding region and the S gene show similar patterns in terms of daily dN and dS rates across time, trending towards decrease over time in both MT and WT samples (Figure 3A-B, 3D-E). However, we did not observe a similarly uniform pattern in the dN/dS ratios (Figure 3C, 3F): despite a steady decrease in the dN/dS ratio for RdRp in MT isolates, the dN/dS ratio for RdRp initially increased during days 100-143 and decreased over time in WT isolates. This could be due to the small number of submitted WT isolates in the later stages, particularly between days 174-203 and 204-242 (32 and 2 isolates, respectively). For the S gene, there was a marked increase in the dN/dS ratio in days 100-143, and a slight decrease over the following stages in MT isolates. On the other hand, the dN/dS ratio for the S gene in the WT isolates initially decreased between days 80-99 and days 100-143, but increased dramatically afterwards, which was followed by another decrease in days 204-242. It should be noted that, in WT isolates, non-synonymous and synonymous mutation rates for the RdRp region during days 204-242 and synonymous mutation rate for the S gene during days 174-203 are uniformly zero, and cannot be visualized on the graph due to the -log10 scale.

**Figure 3.**
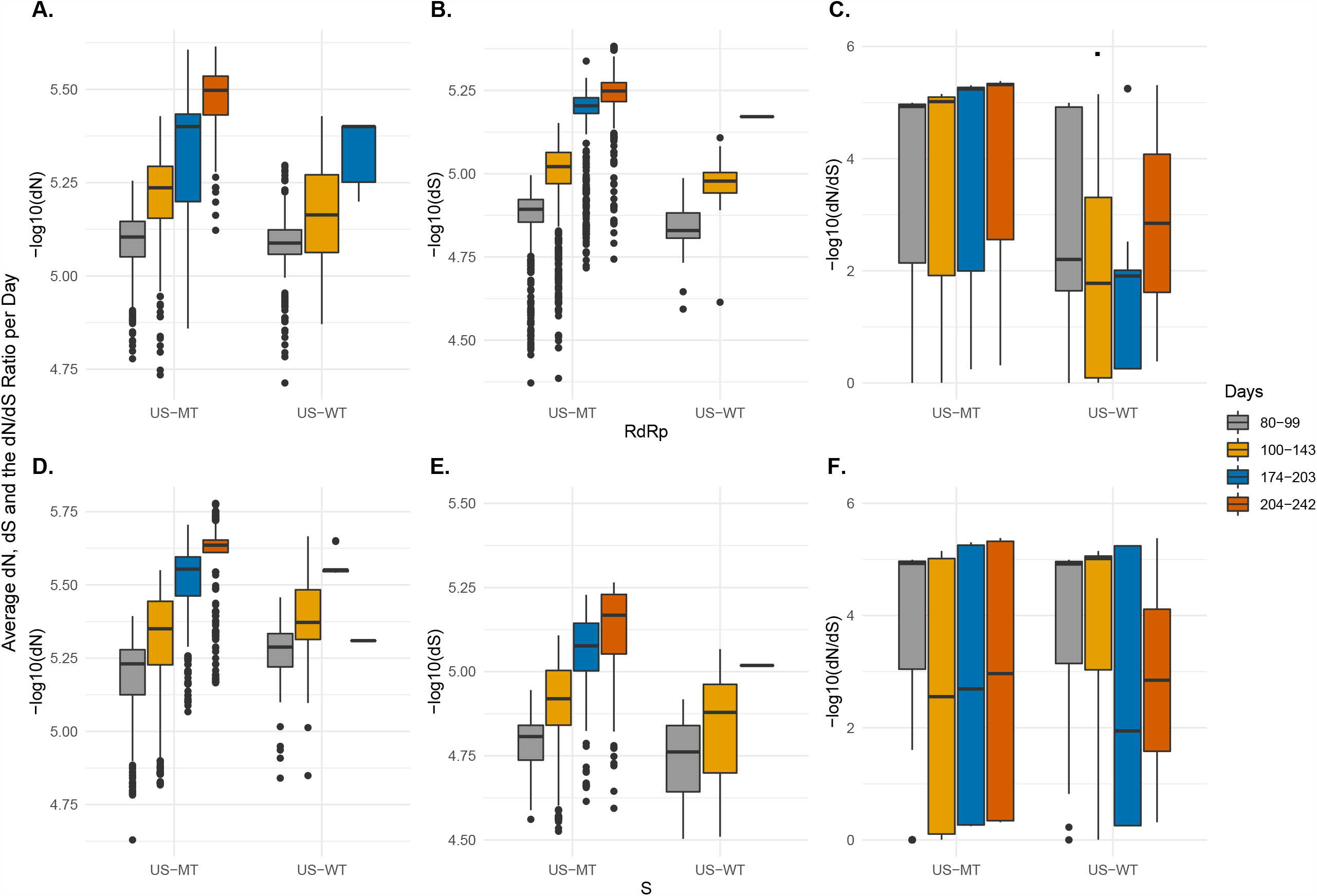
Mutation rates and dN/dS ratios in RdRp and S coding regions per day for country - phenotype pairings of the SARS-CoV-2 isolates from the US divided by pandemic stage. (A-F) Box-and-whiskers plots for mutation rates and dN/dS ratios for the RdRp coding region (A-C) and the S gene (D-F). Due to the -log10 scale, higher values on the y-axis indicate lower mutation rates.

To validate the results obtained from the US isolates, we repeated our analyses with the UK isolate sequences. As the UK only had sufficient isolate sequences available for days 80-99 and 100-143, we focused on these two time periods. We calculated daily mutation rates, dN and dS rates, as well as the dN/dS ratios for each gene and genotype. Our results show a significant difference in dN and dS values and the dN/dS ratios of RdRp and S genes between WT and MT isolates for each time period (p<0.001) (Figure 4). For both genotypes, the dN and dS rates decreased significantly in the plateau stage for the RdRp region and the S gene. In addition, the dN/dS ratio for RdRp increased in days 100-143 in both WT and MT genotypes, whereas the dN/dS ratio of the S gene increased in MT isolates but decreased in WT isolates in days 100-143.

**Figure 4.**
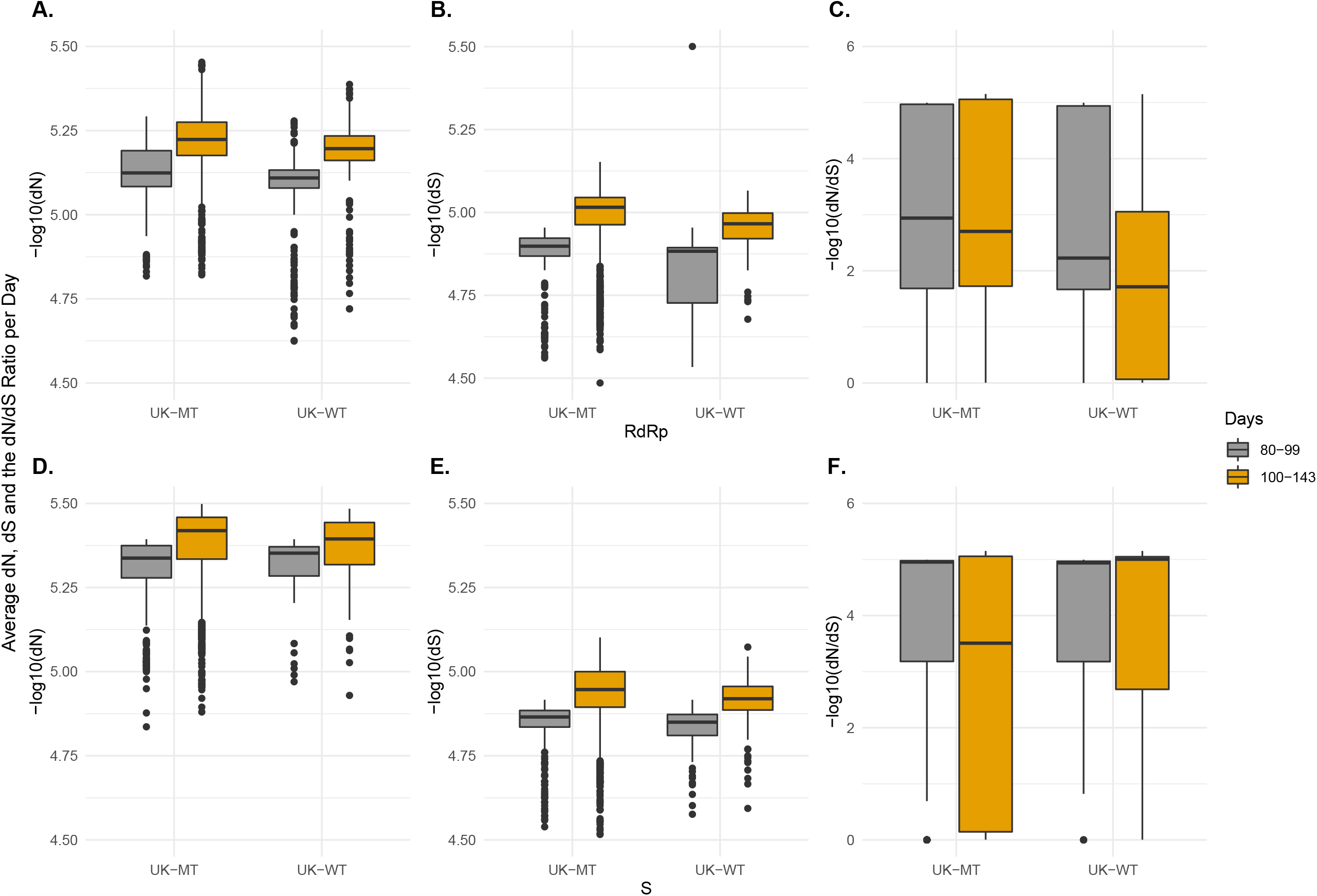
Mutation rates and dN/dS ratios in RdRp and S coding regions per day for country - phenotype pairings of the SARS-CoV-2 isolates from the UK divided by pandemic stage. (A-F) Box-and-whiskers plots for mutation rates and dN/dS ratios for the RdRp coding region (A-C) and the S gene (D-F). Due to the -log10 scale, higher values on the y-axis indicate lower mutation rates.

### Modeling of per site mutation rates indicates that S and RdRp evolve differentially between MT and WT isolates

In addition to the gene-level mutation rate analysis, we next sought to estimate the synonymous and non-synonymous mutation rates per site per day, and the daily dN/dS ratio for each gene and country, using the conservative model previously described by Vega et al. (Supplementary File 1). Our results show that most SARS-CoV-2 genes have undergone negative selection, consistent with previous findings (Berrio, Gartner & Wray, 2020). In both the UK and the US, the dN/dS ratio is highly similar for the majority of genes across genotypes. MT isolates tend to have lower dN/dS ratios for most genes compared to WT isolates. It is noteworthy that, similar to the mutation analysis described above and in Figure 1, RdRp and S are the two most disparate regions between WT and MT isolates in both countries. The dN/dS ratio of RdRp is greater in WT isolates compared to MTs, whereas the opposite is true for the S gene, with its dN/dS ratio being lower in WT isolates compared to MT isolates. It should be noted that Orf8 is the third most disparate gene between MT and WT isolates. The significance of this finding remains to be explored, as there is limited knowledge on the biological function of Orf8 and its possible relationship to the two mutations that define the MT genotype is not clear. However, its proposed role in immune evasion could possibly provide a plausible explanation.

### Mutational dynamics in RdRp and S over time: evolving in opposite directions

As the RdRp region and the S gene were the two genomic regions with the most different dN/dS ratios between MT and WT isolates, we modeled their mutation rates per site in the US for the four time periods described in Figure 3 (Figure 5, Supplementary File 2).

**Figure 5.**
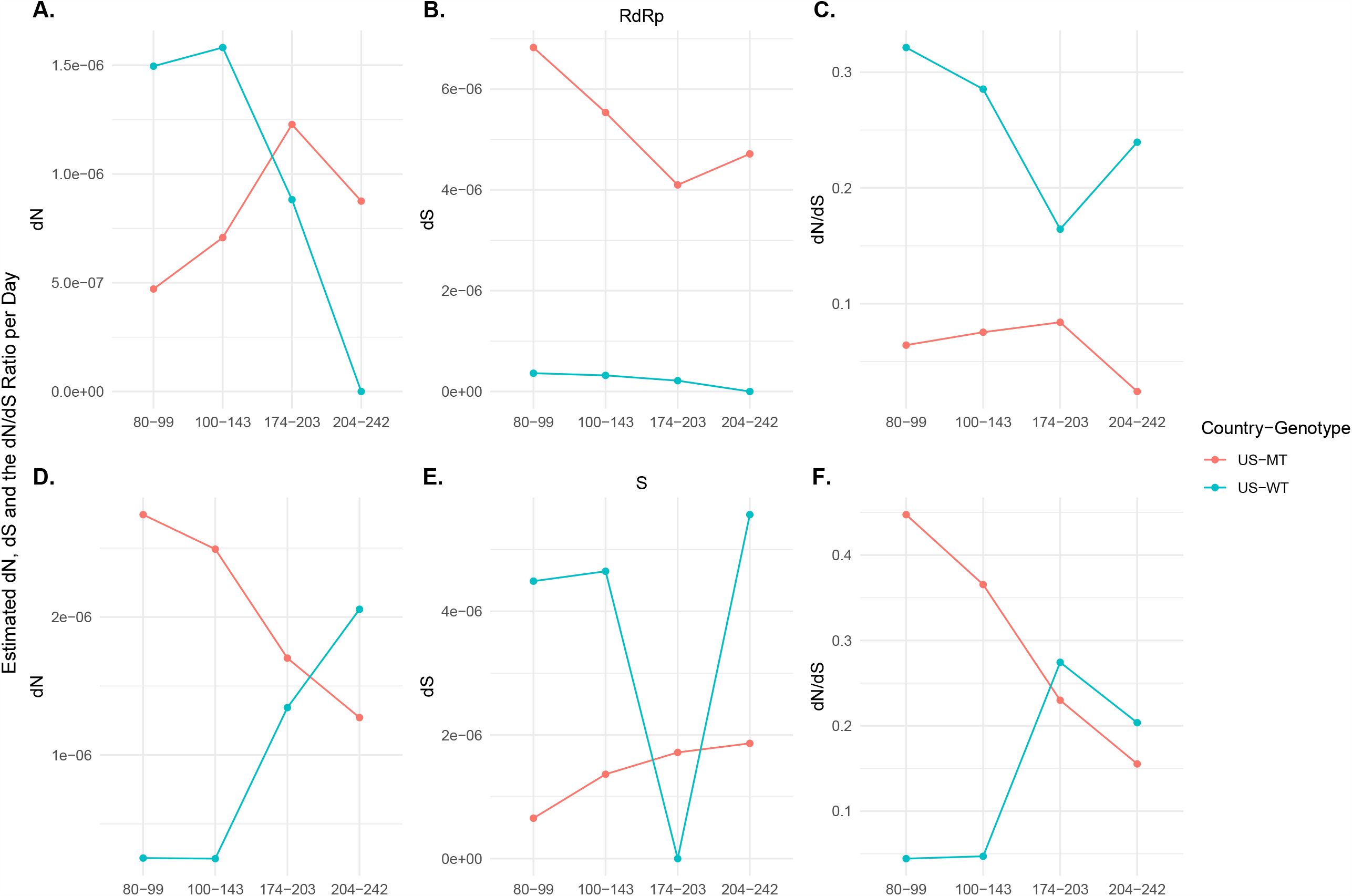
Estimated mutation rates in RdRp and S coding regions per site per day and dN/dS ratios per day in RdRp and S coding regions for country - phenotype pairings of the SARS-CoV-2 isolates from the US divided by pandemic stage. (A-F) Line graphs for estimated mutation rates and estimated dN/dS ratios for the RdRp coding region (A-C) and the S gene (D-F). The estimated values were calculated using the least square fitting.

Our results show that the dN rate of the RdRp coding region in WT isolates slightly increased during days 100-143 and decreased during days 174-203 and 204-242, whereas in MT isolates it increased during days 80-143 and days 174-203, and decreased during days 204-242. We observed that the dS value for the RdRp region decreased over time in WT isolates. MT isolates also showed a similar pattern, except during days 204-242, where the dS value increased instead. While the dN/dS ratio for the RdRp region increased in MT isolates in days 80-143 and days 174-203, it decreased in WT isolates in the same time categories.

For the S gene, the trends were reversed compared to the RdRp. The dN value of the S gene constantly increased in WT isolates but decreased in MT isolates. On the other hand, the dS rate for the S gene increased across time in MT isolates, while there was no consistent pattern for the WT isolates. The sharp decrease in the synonymous mutation rate observed during days 174-203 in WT isolates is likely due to the relatively small number of WT isolates during this period, and should be interpreted with caution. Temporal changes in the dN/dS ratios were similar to the dN rate changes, where it constantly decreased across time periods in MT isolates. Although not as clear as it is in MT isolates, we observed a trend in the opposite direction for the dN/dS ratios in the WT isolates: it initially increased in days 100-143, followed by another sharp increase in days 174-203 and slightly decreased during days 204-242, but the model was not significant for dN/dS for this time period.

## 5. Discussion

Spike mutation D614G has been under spotlight since May 2020 and now it is widely accepted as causing a more transmissible form of SARS-CoV-2. Less is known about its co-mutation in RdRp (P323L), however, it could be leading to altered mutational fidelity. We previously showed that the mutant viruses accumulate mutations differently compared to those without. In this study, we show that viruses with the dominant 23403A>G (S D614G) and 14408C>T (RdRp P323L) mutations also have lower dN/dS ratios compared to those without these two mutations, particularly at RdRp coding region and Orf8 gene. On the contrary, S gene, unlike the other genes, has higher dN/dS ratios in the mutant genomes.

Moreover, temporal analyses point to opposite trends of selection for these two critical genes. While the S gene seems to be under stronger negative selection in WT genomes during the early stages of the pandemic, it is almost at equal levels between MT and WT genomes in the later stages. On the other hand, RdRp is under stronger overall negative selection in the MT genomes, particularly during the early stages. One could speculate that RdRp with the P323L (14408C>T) mutation in the MT genomes has been relatively fit, and did not undergo significant changes in its dN/dS ratio over time. On the other hand, as the pandemic progressed, S with the dominant D614G (23403A>G) mutation has become increasingly less tolerant to non-synonymous mutations. As there are no other frequently observed mutations in the S gene, this dramatic change can be explained by the presence of more limiting epidemiological conditions for the virus in the later stages that led to stronger negative selection over time. It should be noted that these two genes harbor two of the most frequently observed mutations in the SARS-CoV-2 genome, and it is likely that non-synonymous mutations in these two genes/gene regions have relatively greater impact on viral fitness. One question that remains to be answered is whether the RdRp 14408C>T mutation has any effect on differential mutation rates observed between the S gene and the RdRp coding region of the two genotypes. Finally, the underlying reason for the observed association between the dN/dS ratio of the Orf8 gene and the MT genotype remains to be investigated.

## 6. Conclusions

Overall, our results point to S and RdRp as critical players of SARS-CoV-2 evolution. A better understanding of the molecular basis of the evolutionary differences between these two genes/gene regions and the rest of the genome, as well as the effects of S D614G and RdRp P323L mutations, could lead to more effective strategies in combating COVID-19.

## Data Availability

The data is available at Mendeley: Kochan, Necla; Eskier, Doga; Suner, Asli; Karakulah, Gokhan; Oktay, Yavuz (2020), SARS-CoV-2 GISAID UK-US isolates (2020-09-07) genotyping VCF, Mendeley Data, V1, doi: 10.17632/5dfj2hhnng.1

https://10.17632/5dfj2hhnng.1

## Acknowledgements

The authors would like to thank Mr. Aliriza Aribaş from Izmir Biomedicine and Genome Center for his technical assistance. The authors also would like to extend their thanks to Izmir Biomedicine and Genome Center (IBG) COVID19 platform IBG-COVID19 for their support in implementing the study.

## Grant Disclosures

The following grant information was disclosed by the authors:

The Scientific and Technological Research Council of Turkey (TUBITAK-STAR) and Turkish Academy of Sciences Young Investigator Program (TU□BA-GEBI□P).

## Author Contributions

Necla Koçhan, Dog□a Eskier, Asli Suner, Go□khan Karaku□lah and Yavuz Oktay conceived and designed the experiments, performed the experiments, analyzed the data, prepared figures and/or tables, authored or reviewed drafts of the paper, and approved the final draft.

## Data Availability

The data is available at Mendeley: Koçhan, Necla; Eskier, Doğa; Suner, Asli; Karakülah, Gökhan; Oktay, Yavuz (2020), “SARS-CoV-2 GISAID UK-US isolates (2020-09-07) genotyping VCF”, Mendeley Data, V1, doi: 10.17632/5dfj2hhnng.1

## Supplemental Information

Supplemental information for this article is available.

